# Aggregate expression of genes lacking CpG islands increases with age and is positively associated with human mortality

**DOI:** 10.1101/2022.09.03.22279290

**Authors:** Richard A. Kerber, Elizabeth O’Brien, Richard M. Cawthon

## Abstract

In both mice and humans the misexpression of many genes lacking CpG islands (CGI- genes) increases with age, promoting inflammation and degenerative changes (Lee et al. 2021, ref. 1). In light of this recent discovery, we have revisited and expanded upon our previous work on gene expressions vs. aging and mortality in the three-generation CEPH (Centre d’Etudes du Polymorphisme Humain) Utah (CEU) families (Kerber et al. 2009, ref. 2). That study examined gene expressions in lymphoblastoid cell lines (LCLs) established in the early 1980s from all three CEU generations, in relation to age at blood draw, and in relation to the long-term survival of the grandparent generation. The 2009 study did not, however, consider the CGI status of genes, and it excluded from analysis genes not expressed in all of the subjects; therefore, the contribution to variation in age-at-death of inter-individual variation in the misexpression of genes with increasing age was not investigated. For the current study, after categorizing genes by their CGI status (- or +), we now find that most CGI- gene expressions in the LCLs increased with donor age, and after adjustment for donor age and sex, were positively associated with mortality risks. In contrast, most CGI+ gene expressions decreased with donor age, with higher expressions associated with decreased mortality risks. Of 7025 genes with known CGI status with expression detected in sufficient numbers of subjects from the grandparent generation to allow testing of association with mortality, 1834 genes were expressed in all subjects’ LCLs across all three generations, and 5191 were expressed in some, but not all subjects. We found the set of “not always expressed” genes to be highly enriched for CGI- genes. Furthermore, 49.4% of the CGI- genes were never expressed from ages 0-14, but expressed sometimes or always at older ages; in contrast, only 22.3% of the CGI+ genes were never expressed from ages 0-14, but expressed at older ages. These data support the model proposed by Lee et al. 2021, whereby tissue-restricted CGI- gene expressions become increasingly misexpressed during aging, contributing to loss of cellular identity, multiple aging-related pathologies, and ultimately death.

## Introduction

Lee et al. recently showed [1] that the misexpression of many genes lacking CpG islands (CGI-genes) increases with age in both mice and humans, is higher in patients with aging-associated diseases than in healthy age-matched controls, and is suppressed in aged mice receiving various treatments known to extend lifespan. Lee et al. [1] further showed that these CGI-gene misexpressions promote inflammation and the development of degenerative changes, supporting the inflammaging theory of aging [3].

Here we investigate associations of CGI- and CGI+ genes’ expressions with human mortality using gene expression data from lymphoblastoid cell lines (LCLs) established in the early 1980s from the three-generation Utah CEPH (Centre d’Etudes du Polymorphisme Humain) families and available long-term follow-up survival data for the grandparent generation [2].

## Materials and Methods

CEPH ⁄ Utah (CEU) families’ Epstein-Barr-virus-immortalized lymphoblastoid cell lines (LCLs), and DNA extracted therefrom, are available from the Coriell Cell Repositories in Camden, NJ, USA (http://ccr.coriell.org/Sections/Collections/NIGMS/CEPHFamilies.aspx?PgId=49&Coll=GM), for 46 three-generation CEU families, each pedigree consisting of 5–15 siblings, their two parents, and two to four grandparents who were alive when blood samples were collected in the early 1980s [2]. The cell lines include 153 from grandparents. Approximately 20 years of follow-up survival and mortality data are available from the Utah Population Database (UPDB) and the Utah Genetic Reference Project at the University of Utah for 145 of the 153 grandparents [2].

Gene expression levels in many of the CEU cell lines have been determined by several laboratories and made available online to researchers [2]. It is now well established that inherited genetic differences among individuals determine much of the variation in expression levels in these cell lines.

After applying several inclusion/exclusion criteria [2], expression levels for 8174 genes were available from 238 CEU family members across three generations, including 104 grandparents for whom survival data were available; some genes were found to be expressed in all of the subjects’ LCLs, other genes were expressed in only some subjects’ LCLs. CGI status (- or +) from reference 1 was available for 7025 of the 8174 genes.

### Environmental exposures

While inter-individual differences in gene expression are largely attributable to inherited genetic variation, these differences in expression may also reflect environmental exposures (e.g. cigarette smoke) occuring at any time prior to blood draw. Including in our analyses data available from the UPDB on CEU family members’ affiliations with the Church of Jesus Christ of Latter-day Saints (LDS church), and known rates of smoking and non-smoking for LDS vs. non-LDS women and men, we expect only a small number (probably less than ten) of the grandparents to have been smokers [2]. Reduced smoking and alcohol consumption among active church members probably accounts for 1.3 additional years of life expectancy compared to Utahans unaffiliated or inactive in the church [2].

Spousal gene expression profiles were found to be slightly more strongly correlated than expected by chance [2]. However, the correlation of mortality risk between spouses was only 0.075 (P-value 0.45), so correlations in gene expression are not likely to confound the survival analysis.

### Age at blood draw analyses

Gene expression as a function of age at draw was analyzed in two ways. Treating the grandparents’ expression data as independent observations, we used ordinary least squares methods to regress expression level for each of the always-expressed probesets against age, adjusting for sex.

Since gene expressions show familial correlations, standard linear regression approaches are not appropriate for the study of age-related variation in expression patterns in sets of closely related subjects. Therefore, we used linear mixed-effects models to adjust for the kinship among family members. We considered both linear and quadratic effects for age at draw in these three-generation families. As for the grandparents only analysis, expression level was modeled as a function of age at draw, age squared, and sex. Heritability estimates are available in reference 2.

### Mortality models

In the grandparent generation age at blood draw ranged from 57 to 97 years of age. Median follow-up age was 84.7 years (range 65.7–100.8). Survival was measured from age at blood draw to age at death or age last known alive. Proportional hazards models adjusted for sex, birth year, and age at blood draw were used to test the association of each gene’s expression level with mortality. Nonproportionality for each proportional hazards model was tested using the cox.zph function in the R software package (http://www.r-project.org). Some nonproportionality was found with P-values < 0.0001; however, none of the genes strongly associated with mortality had a P-value for nonproportionality less than 0.28 (EMP3).

### Bivariate age-at-draw vs. survival models, and adjustment for multiple comparisons

Our procedures for testing the composite null hypothesis that gene expression was related to neither aging nor longevity, and for adjusting for multiple testing, are described in detail in reference 2. We employed a simple Bonferroni correction for the univariate analyses, and a Monte Carlo permutation test for the bivariate analyses.

Two-sample Wilcoxon tests were used to compare the distribution of Z-scores for age and longevity between CGI- and CGI+ genes.

## Results

Table 1 summarizes age-related change and mortality Z-score results for the full dataset of 7025 genes that are provided in Supplementary Table 1. A positive age-related change Z-score means that in these cross-sectional data, the expression level of the gene increased with increasing age of the blood donors; and a negative Z-score means expression decreased with increasing age of donor. A positive mortality Z-score means that in these cross-sectional data, higher expression of the gene was associated with higher mortality; and a negative mortality Z-score means that higher expression was associated with lower mortality.

**Table 1.**
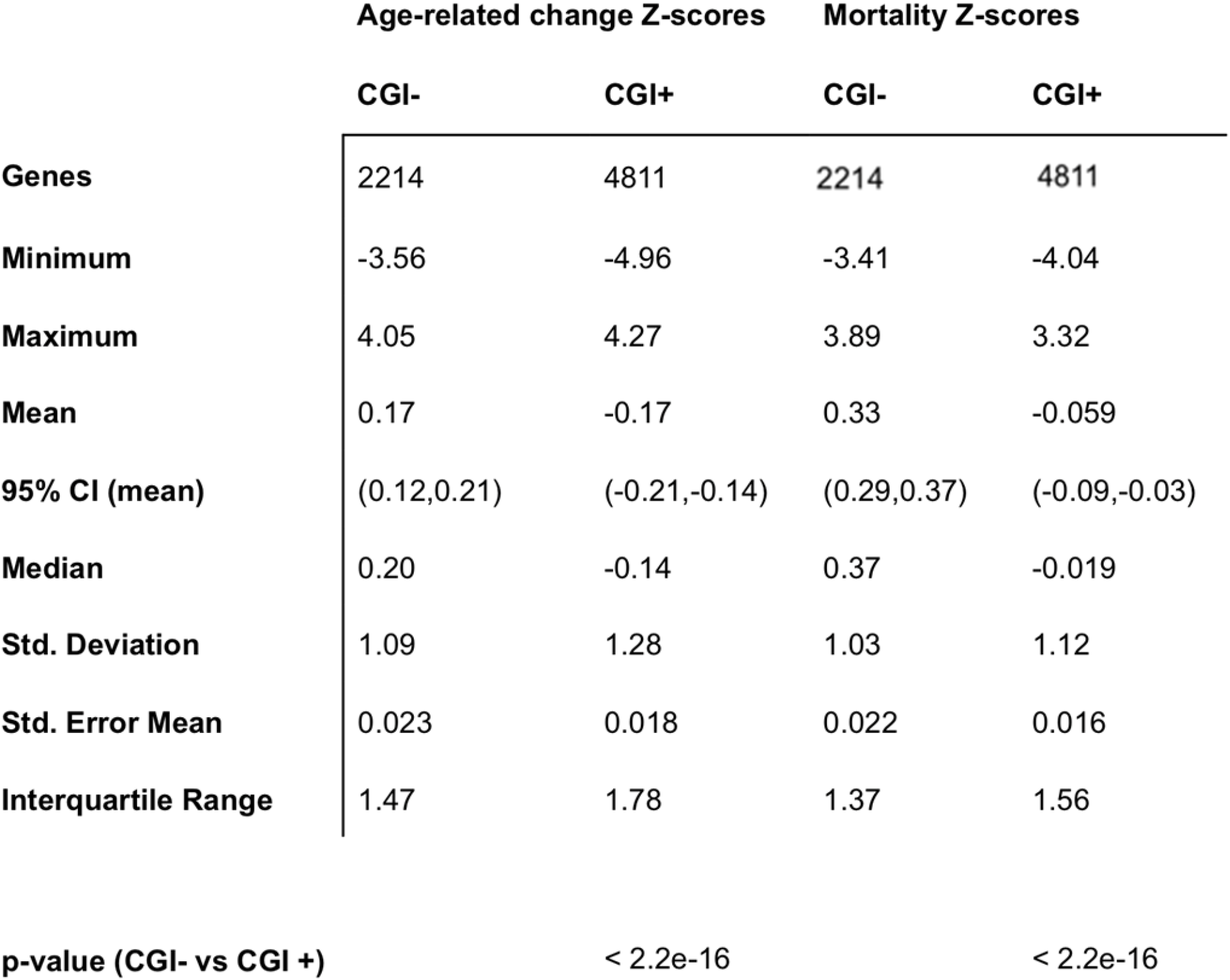
Associations of CGI- and CGI+ gene expressions with aging and mortality.

The scatterplot below depicts the age-related change and mortality Z-scores for the 2214 CGI-genes (red dots), and 4811 CGI+ genes (black dots).

**Figure 1.**
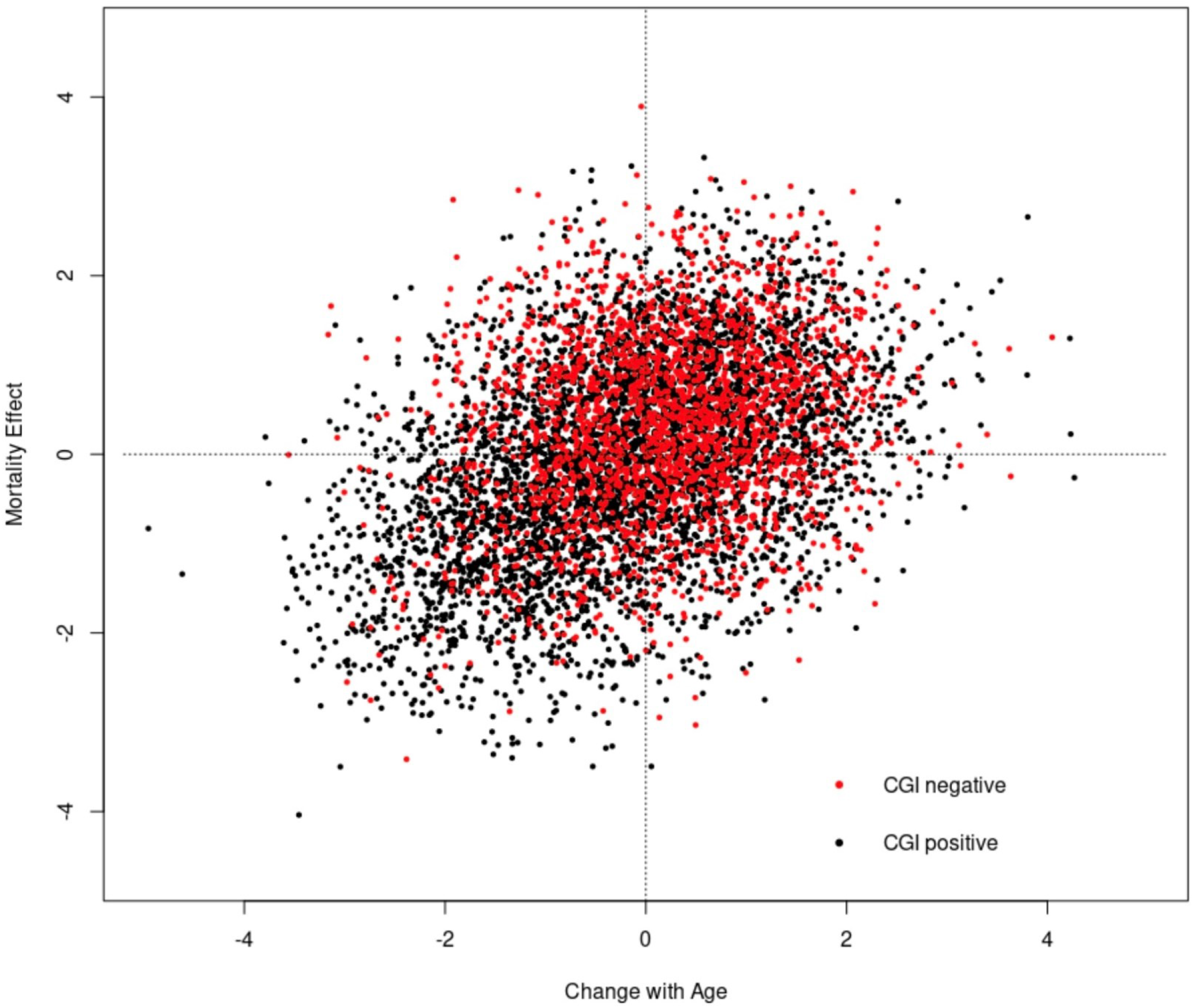
Change with age vs. mortality for 7025 gene expressions in Utah CEPH lymphoblastoid cell lines.

In our previous study of gene expressions vs. aging and mortality in these families [2], we included only gene expressions that were detected in all subjects from all three generations for whom expression data were available, i.e. “always expressed genes”. For the present analysis of genes with known CGI status, we studied 1834 genes that were always expressed, and 5191 additional genes whose expressions were detected in some, but not all subjects across the three generations. We found the “not always expressed” set of genes to be highly enriched for CGI-genes. We found that 49.9% of CGI-genes were never expressed in the cell lines from donors aged 0-14 years, but sometimes or always expressed in the cell lines from older donors (Table 2 and Figure 2); in contrast, 22.4% of CGI+ genes were never expressed in cell lines from donors aged 0-14 years, but sometimes or always expressed in the cell lines from older donors.

**Table 2.** Genes first expressed after childhood.

| CGI Status | Total genes | "Never expressed" status is lost at ages > 14 |  |
| --- | --- | --- | --- |
| Negative | 2214 | 1105 | 49.9% |
| Positive | 4811 | 1076 | 22.4% |

**Figure 2.**
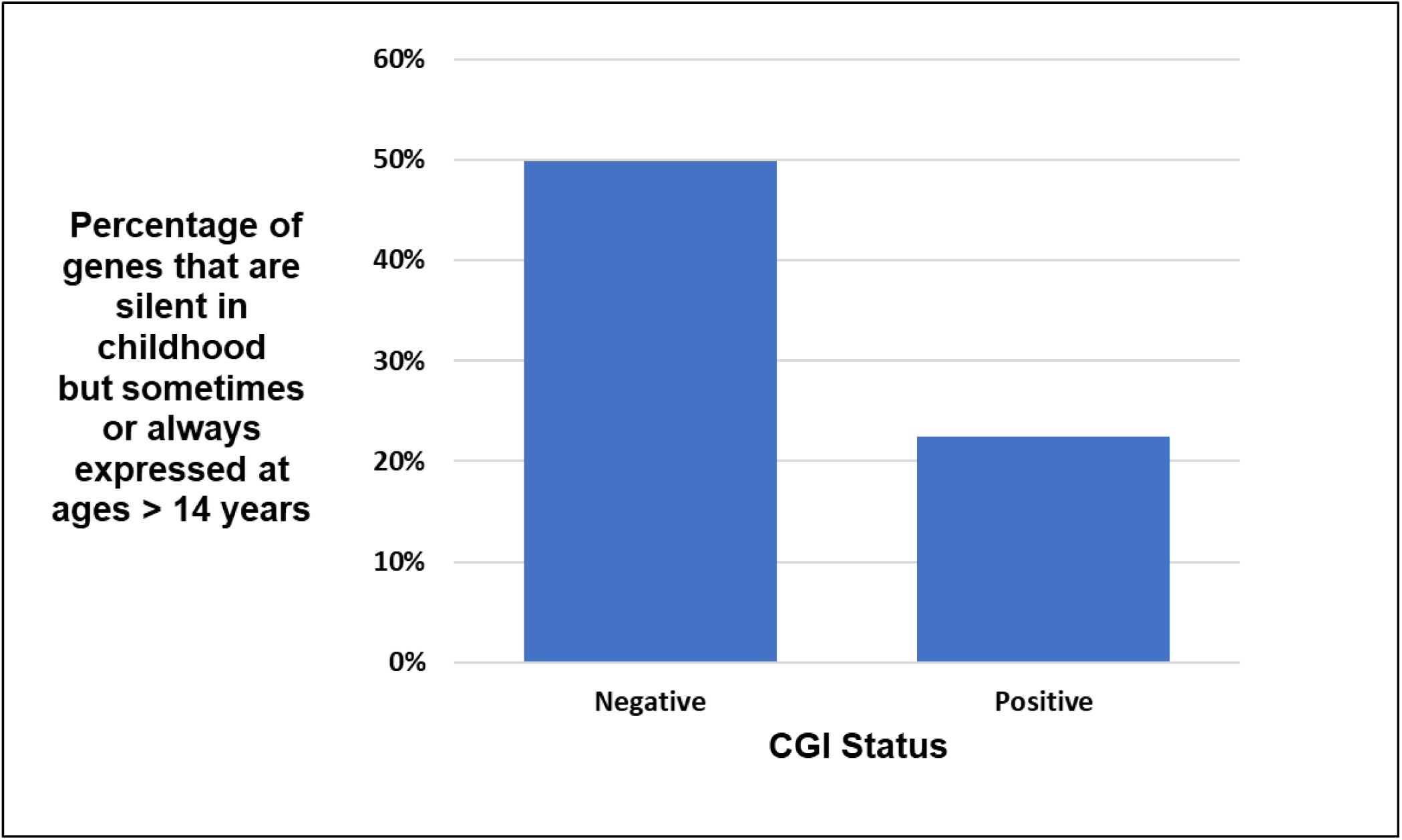
Loss of gene silencing after age 14 by CGl status.

## Conclusions

These data support the model proposed by Lee et al. [1], whereby tissue-restricted CGI-gene expressions become increasingly misexpressed during aging, contributing to loss of cellular identity, multiple aging-related pathologies, and ultimately death. Future investigations may identify a subset of CGI-gene misexpressions that can serve as an accurate and convenient measure of the rate of senescent aging at a single timepoint in adults. As has been pointed out in a recent review [4], reliable “aging clocks” should eventually prove useful in clinical medicine, allowing the identification of the patients at highest risk of aging-related disabilities and diseases well before the onset of those pathologies. Furthermore, repeatedly and longitudinally testing the rate of the aging clock in cohorts of rapidly aging subjects in clinical trials may soon identify safe and effective behavioral and pharmacologic interventions to slow senescent aging.

## Data Availability

All data produced in the present study are available upon reasonable request to the authors.

